# SARS-CoV-2 and human milk: what is the evidence?

**DOI:** 10.1101/2020.04.07.20056812

**Authors:** Kimberly A. Lackey, Ryan M. Pace, Janet E. Williams, Lars Bode, Sharon M. Donovan, Kirsi M. Järvinen, Antti E. Seppo, Daniel J. Raiten, Courtney L. Meehan, Mark A. McGuire, Michelle K. McGuire

## Abstract

The novel coronavirus SARS-CoV-2 has emerged as one of the most compelling and concerning public health challenges of our time. To address the myriad issues generated by this pandemic, an interdisciplinary breadth of research, clinical, and public health communities has rapidly engaged to collectively find answers and solutions. One area of active inquiry is understanding the mode(s) of SARS-CoV-2 transmission. While respiratory droplets are a known mechanism of transmission, other mechanisms are likely. Of particular importance to global health is the possibility of vertical transmission from infected mothers to infants through breastfeeding or consumption of human milk. However, there is limited published literature related to vertical transmission of any human coronaviruses (including SARS-CoV-2) via human milk and/or breastfeeding. Results of the literature search reported here (finalized on April 17, 2020) revealed a single study providing some evidence of vertical transmission of human coronavirus 229E; a single study evaluating presence of SARS-CoV in human milk (it was negative); and no published data on MERS-CoV and human milk. We identified 12 studies reporting human milk tested for SARS-CoV-2; one study detected the virus in one milk sample, and another study detected SARS-CoV-2 specific IgG in milk. Importantly, none of the studies on coronaviruses and human milk report validation of their collection and analytical methods for use in human milk. These reports are evaluated here, and their implications related to the possibility of vertical transmission of coronaviruses (in particular, SARS-CoV-2) during breastfeeding are discussed.

## INTRODUCTION

The global pandemic caused by the SARS-CoV-2 virus is one of the most compelling and concerning global health crises of our time. Fortunately, this pandemic has rapidly mobilized the full range of expertise represented by researchers, clinicians, and public health officials. While our understanding of the biology, clinical implications, and strategies for mitigation continues to evolve, one issue that has received limited attention is the implication of this pandemic for infant feeding practices. This lack of attention has resulted in mixed messages regarding guidance about optimal infant feeding practices (e.g., American Academy of Pediatrics, 2020; Centers for Disease Control and Prevention, 2020a; World Health Organization, 2020a; United Nations Children’s Fund, 2020) and a consequent lack of confidence about best approaches to infant feeding in the face of this growing pandemic. Even when a mother is positive for COVID-19, the World Health Organization (WHO) recommends breastfeeding be initiated within 1 hr of birth, exclusive breastfeeding be continued for 6 mo, and breastfeeding be continued for up to 2 years. They suggest use of appropriate respiratory hygiene, hand hygiene, and environmental cleaning precautions. The United Nations Children’s Fund (UNICEF) recommends that COVID-19-positive mothers continue breastfeeding while applying precautions, such as wearing a mask and handwashing before and after feeding (United Nations Children’s Fund, 2020). The US Centers for Disease Control and Prevention (CDC) neither recommends nor discourages breastfeeding but advises that decisions be made by the mother and family in consultation with their healthcare providers (Centers for Disease Control and Prevention, 2020a). They recommend that during temporary separation (should that occur) mothers who intend to breastfeed should express their milk using proper hand hygiene, and that the expressed milk should be fed to the newborn by a healthy caregiver. Further, if a mother and newborn do room-in and the mother wishes to feed at the breast, the CDC recommends that she should wear a facemask and practice hand hygiene before each feeding.

### Box 1

**KEY MESSAGES**

- Very little is known about coronaviruses in human milk and whether brxeastfeeding is a possible mode of vertical transmission.
- Limited, weak evidence suggests that some coronaviruses (including SARS-CoV-2) may be present in human milk, but these studies do not report methods of sample collection and validation of RT-PCR assays for human milk.
- Nothing is known about the timing of the antibody response in human milk to SARS-CoV-2 infection.
- Future research should utilize validated methods and focus on both potential risks and protective effects of breastfeeding.

It is well established that viral transmission through human milk can occur (Jones, 2001; Lawrence & Lawrence, 2004). Notable examples include human immunodeficiency virus (HIV; Black, 1996; Ziegler, Johnson, Cooper, & Gold, 1985), cytomegalovirus (CMV; Stagno & Cloud, 1994), and human T-cell lymphotropic virus type 1 (HTLV-1; Boostani, Sadeghi, Sabouri, & Ghabeli-Juibary, 2018). Perhaps the most prominent example of mother-to-child viral transmission via breastfeeding is HIV infection, during which higher milk and serum viral loads are associated with an increased risk of transmission (Davis et al., 2016; Semba et al., 1999; Willumsen et al., 2003). The risk of postnatal infection for breastfed infants of HIV+ mothers is ≈10-20% over the first 2 years of life without the use of antiretroviral therapies (ART; Dunn, Newell, Ades, & Peckham, 1992; Nduati et al., 2001). However, compared to mixed feeding, exclusive breastfeeding is associated with lower risk of transmission of HIV infection to infants (Coutsoudis et al., 2001; Iliff et al., 2005). In many high-income nations, breastfeeding is contraindicated in the case of maternal HIV infection with or without maternal ART (e.g., American Academy of Pediatrics, 2012; Centers for Disease Control and Prevention, 2020b). Conversely, in low-and-middle-income nations, infant mortality from malnutrition and infectious disease may outweigh the risk of acquiring HIV via vertical transmission during breastfeeding. As such, breastfeeding is recommended (World Health Organization, 2016).

With respect to CMV, it is estimated that 60-70% of breastfed infants of CMV-seropositive mothers become infected with CMV (Dworsky, Yow, Stagno, Pass, & Alford, 1983; Minamishima et al., 1994). The risk of CMV infection in neonates is highest in preterm and very low birthweight (<1500 g) infants (Hamprecht & Goelz, 2017; Lanzieri, Dollard, Josephson, Schmid, & Bialek, 2013). A small percentage of infected infants develop a severe complication known as CMV sepsis-like syndrome, which can be fatal (Fischer et al., 2010). Nonetheless, breastfeeding is not contraindicated in CMV-seropositive women with healthy, term infants (American Academy of Pediatrics, 2012; Centers for Disease Control and Prevention, 2019b; World Health Organization, 2009).

For HTLV-1, breastfeeding is considered the major route of infection for infants (Moriuchi, Masuzaki, Doi, & Katamine, 2013). HTLV-1 infection is lifelong and, while most infected individuals remain asymptomatic, approximately 10% develop severe disease, including adult T-cell leukemia, a highly aggressive and usually fatal malignancy (Rosadas & Taylor, 2019). Some organizations and agencies list maternal HTLV-1 as a contraindication for breastfeeding (American Academy of Pediatrics, 2012; Centers for Disease Control and Prevention, 2019a), while others do not (World Health Organization, 2009).

Human coronaviruses are enveloped, positive-sense, single-stranded RNA viruses first described in 1965 (Tyrrell & Bynoe, 1965). There are 7 identified strains known to infect humans. Four of the strains (alphacoronaviruses 229E, NL63, and OC43; betacoronavirus HKU1) are ubiquitous in humans and cause the common cold. There is limited evidence that one of these (229E) may be vertically transmitted from mothers to infants, although the mechanism remains unclear (Gagneur et al., 2008). The presence of 229E in neonatal gastric samples suggests that one possible mechanism for infection is through human milk, although this study did not evaluate human milk specifically (Gagneur et al., 2008).

In light of the emergence of the novel coronavirus SARS-CoV-2, several issues related to human milk and coronavirus infection demand immediate attention, the first and foremost being whether or not the virus is present in human milk produced by infected or exposed women. Of particular interest in this context are 1) the potential role that breastfeeding could play in vertical transmission of SARS-CoV-2 from women to infants via human milk; and 2) the potential protective effects of targeted antibodies and other immunoprotective components in human milk against COVID-19. The goal of this review was to evaluate the published evidence regarding the presence of this and other human coronaviruses in human milk.

## METHODS

We used a variety of databases to identify relevant literature published as of April 17, 2020, and the list of databases and search terms used can be found in **Table 1**. It is noteworthy that in addition to using standard scientific databases (e.g., PubMed) we also used a general Google search and a search of preprint servers to identify reports that had not yet been published in refereed journals (i.e., gray literature). Any research in which human milk was collected and tested for a human coronavirus was included in this review.

**Table 1.**
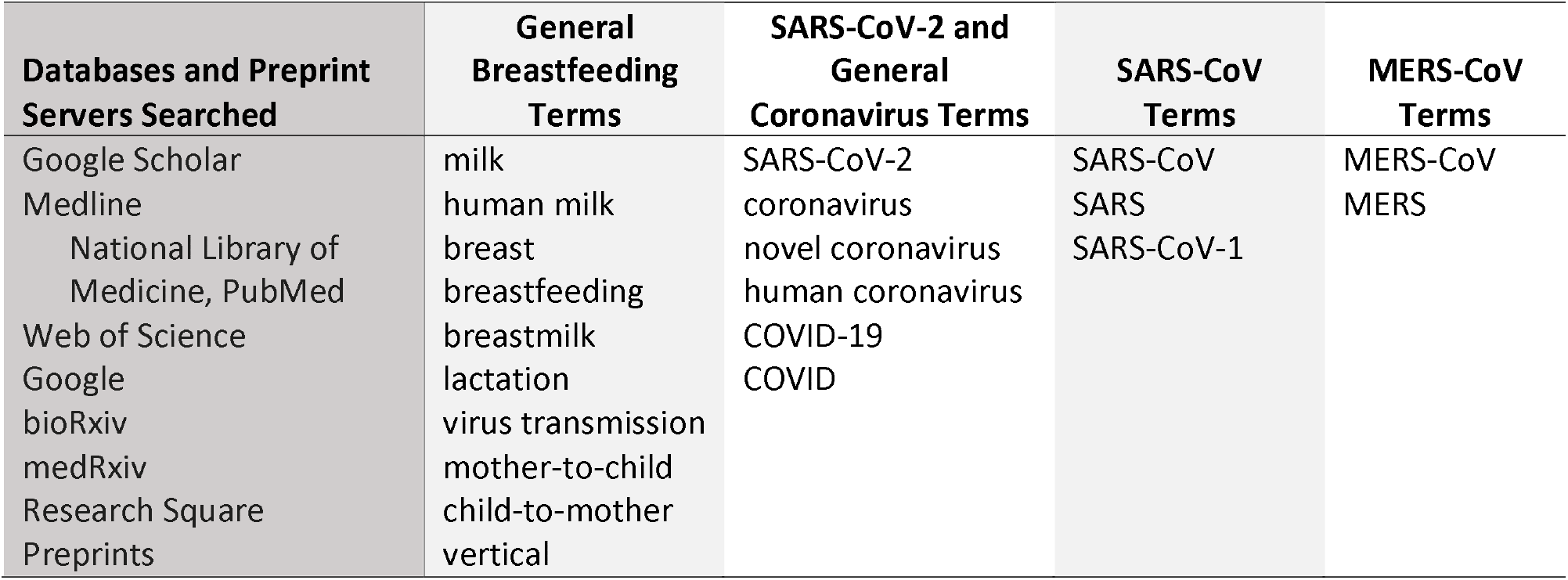
Search terms, databases, and preprint servers used to identify existing literature reporting the possibility of vertical transmission of coronaviruses from mother to infant during breastfeeding as of April 17, 2020.

## RESULTS

### MERS-CoV

The deadliest of the human coronaviruses to date is MERS-CoV, which emerged in Saudi Arabia in 2012. The disease caused by MERS-CoV, Middle Eastern respiratory syndrome (MERS), is characterized by severe respiratory illness with symptoms of fever, cough, and shortness of breath. MERS-CoV is a betacoronavirus, and the case fatality rate of MERS is 34% (Mahase, 2020). There are no reports of the presence or absence of MERS-CoV in human milk. However, there are reports of the presence of MERS-CoV in the milk of dromedary camels (*Camelus dromedaries*; Conzade et al., 2018; Hemida et al., 2015; Reusken et al., 2014), and there is one report of a human likely infected through the consumption of raw (unpasteurized) camel milk (Memish et al., 2014). In camel milk samples spiked with MERS-CoV, viable virus could still be recovered after 48 hr (van Doremalen, Bushmaker, Karesh, & Munster, 2014). These observations resulted in recommendations against consuming raw, unpasteurized camel milk (World Health Organization, 2019). It is unclear if there is vertical transmission of MERS-CoV between camelid cows and their calves, and whether infection occurs as a direct result of lactation/nursing in this species. There are no data on vertical transmission of MERS-CoV between women and their infants (Jeong et al., 2017; Schwartz & Graham, 2020).

### SARS-CoV

A related virus, SARS-CoV, emerged in 2003 in China, although the disease (severe acute respiratory syndrome, SARS) quickly spread globally. SARS is clinically manifested by fever, dry cough, headache, muscle aches, and difficulty breathing. No treatment exists except supportive care, but there have been no reports of SARS-CoV transmission since 2004. Like MERS-CoV, SARS-CoV is a betacoronavirus, and the case fatality rate of SARS is estimated at 10% (Mahase, 2020).

Currently, there is one report in which human milk was tested for SARS-CoV (Robertson et al., 2004), and two reports of human milk being tested for SARS-CoV antibodies (Robertson et al., 2004; Stockman, Lowther, Coy, Saw, & Parashar, 2004). Robertson and colleagues described a woman infected during the second trimester of pregnancy (19 wk). A single milk sample was collected 131 days after the onset of symptoms, but no additional detail on the collection methodologies was provided. Milk was submitted to the CDC, where it was analyzed using reverse transcription polymerase chain reaction (RT-PCR) for viral nucleic acids, and enzyme immunoassay and indirect immunofluorescence to evaluate antibody presence. No additional details on analytical methods were provided. While no viral RNA was detected, antibodies to SARS-CoV were identified in the milk. The infant in this study was never tested for SARS-CoV infection. Stockman and colleagues described a 38-yr-old woman infected in the first trimester of pregnancy (7 wk). She recovered fully and delivered a healthy male infant at 36 wk of gestation. Milk samples were collected at 12 and 30 d postpartum and tested for SARS-CoV antibodies; all were negative. No details on the collection or analysis of the milk were provided. The infant in this study tested negative for SARS-CoV. In both these studies, it is possible that the women had stopped shedding the virus before the milk samples were collected. SARS-CoV shedding in other biological samples typically peaks 12-14 d after the onset of disease (Cheng et al., 2004). There are no documented cases of vertical transmission of SARS-CoV between mothers and infants (Schwartz & Graham, 2020).

### SARS-CoV-2

The novel coronavirus SARS-CoV-2 was named after SARS-CoV due to its shared sequence homology (77.9%; Kim et al., 2020) and similar clinical characteristics. The first reported cases of SARS-CoV-2 infection emerged in late 2019 in China. While the current estimated case fatality rate for COVID-19 (the disease caused by the SARS-CoV-2 virus) is much lower than those of SARS and MERS at roughly 2% (Mahase, 2020), the spread of this pathogen has been much more rapid and extensive.

At the time of writing, there were 12 studies (7 case studies and 5 case series; 3 of which were preprints, or preliminary reports that had not been formally peer-reviewed; **Table 2**) reporting direct testing of milk produced by women infected with SARS-CoV-2 (Chen et al., 2020; Dong et al., 2020; Fan et al., 2020; Y. Li et al., 2020; Weiyong Liu et al., 2020a; Wei Liu et al., 2020b; Wang et al., 2020; Yu et al., 2020; Wu et al., 2020) or by women whose infants were infected (Cui et al., 2020; Kam et al., 2020; Yuehua et al., 2020). In total, 46 milk samples produced by 30 women had been tested; all but one sample (Wu et al., 2020; non-peer-reviewed preprint) were negative for the presence of the virus. Two milk samples produced by a single woman were tested for SARS-CoV-2 specific antibodies; IgG but not IgM was identified in both samples (Yu et al., 2020; non-peer-reviewed preprint). A description of the relevant characteristics for the women and infants in these studies can be found in **Table 2**. Investigators conducting 8 of the 12 studies analyzed milk samples collected at birth or shortly thereafter, reporting only findings in colostrum or transitional milk. Those same eight studies reported on the milk produced by women who were infected during the third trimester of pregnancy, while the other four reported findings from milk produced by mothers of infants infected at 1.5 mo, 3 mo, 6 mo, and 13 mo of age (Cui et al., 2020; Kam et al., 2020; Yuehua et al., 2020, Yu et al., 2020). For the infants born to women infected during pregnancy, most were immediately separated from their mothers post-delivery and were not breastfed for the duration of the period observed in their respective reports. Fourteen of the 30 infants described in these reports were born via cesarean section; only two were specified as vaginal births. Repeated milk samples, collected up to 27 days apart, were analyzed for 8 of the women. All the studies were conducted in China (Chen et al., 2020; Cui et al., 2020; Dong et al., 2020; Fan et al., 2020; Y. Li et al., 2020; Weiyong Liu et al., 2020a; Wei Liu et al., 2020b; Wang et al., 2020; Yuehua et al., 2020, Yu et al., 2020; Wu et al., 2020) or Singapore (Kam et al., 2020).

**Table 2.** Characteristics of women and infants for whom human milk has been sampled and tested for SARS-CoV-2 using RT-PCR and for SARS-CoV-2 specific antibodies>.

| *Publication* | Subjects<br>(n) | Location | Repeated<br>samples | Time of milk<br>collection | Maternal<br>age (yr) | Gestational age<br>at time of<br>maternal<br>infection | RT-PCR<br>results | Milk<br>antibody<br>results | Infant age<br>at the time<br>of infant<br>infection | Infant<br>sex | Delivery<br>mode | Infant<br>breastfed |
| --- | --- | --- | --- | --- | --- | --- | --- | --- | --- | --- | --- | --- |
| *Chen et al.,*<br>*2020* | 6<sup>1</sup> | China | no | d 1 | 27 | 38 wk, 2 d<sup>2</sup> | – | NA | NA | NS | cesarean | NS |
|  |  |  |  |  | 26 | 36 wk, 2 d<sup>2</sup> | – |  | NA | NS | cesarean | NS |
|  |  |  |  |  | 26 | 38 wk, 1 d<sup>2</sup> | – |  | NA | NS | cesarean | NS |
|  |  |  |  |  | 26 | 36 wk, 3 d<sup>2</sup> | – |  | NA | NS | cesarean | NS |
|  |  |  |  |  | 28 | 38 wk<sup>2</sup> | – |  | NA | NS | cesarean | NS |
|  |  |  |  |  | 34 | 39 wk, 4 d<sup>2</sup> | – |  | NA | NS | cesarean | NS |
| *Cui et al.,*<br>*2020* | 1 | China | yes | 55-57 d | NS | NA | – | NA | 50 d | female | NS | yes |
| *Dong et al.,*<br>*2020* | 1 | China | no | 6 d | 29 | 34 wk, 2 d | – | NA | NA | female | cesarean | no |
| *Fan et al.,*<br>*2020* | 2 | China | yes | d 1, 17 | 34 | 37 wk | – | NA | NA | female | cesarean | no |
|  |  |  | no | d 1 | 29 | 36 wk | – |  | NA | female | cesarean | no |
| *Kam et al.,*<br>*2020* | 1 | Singapore | no | 6 mo | NS | NA | – | NA | 6 mo | male | NS | yes<sup>3</sup> |
| *Li et al.,*<br>*2020* | 1 | China | yes | d 1, 2, 3 | 30 | 35 wk | – | NA | NA | male | cesarean | NS |
| *Weiyong Liu*<br>*et al.,*<br>*2020a†* | 2<sup>4</sup> | China | yes | d 2, 3, 12 | 34 | 40 wk | – | NA | NA | male | cesarean | no |
|  |  |  | no | d 2 | 30 | 37 wk | – |  | NA | unclear | vaginal | NS |
| *Wei Liu et*<br>*al., 2020b†* | 10<sup>5</sup> | China | no | NS | NS | NS | – | NA | NA | mixed | NS | no |
| *Wang et al.,*<br>*2020* | 1 | China | no | 36 hr | 34 | 40 wk | – | NA | NA | male | cesarean | no |
| *Wu et al.,*<br>*2020†* | 3<sup>6</sup> | China | yes | d 1, 6, 27 | 29 | 35 wk 4 d | – | NA | NA | NS | cesarean | NS |
|  |  |  |  | d 1, 6, 27 | 28 | 35 wk 5 d | – |  | NA | NS | cesarean | NS |
|  |  |  |  | d 1, 3, 6, 27 | 27 | 38 wk 2 d | + |  | NA | NS | vaginal | NS |
| *Yu et al.,*<br>*2020†* | 1 | China | yes | d 1, 8, 15, 18, 24<sup>7</sup> | 32 | NA | –<sup>8</sup> | IgG+,<br>IgM-<sup>9</sup> | 13 mo | male | NS | yes |
| Yuehua et al., 2020 | 1 | China | no | 3 mo | NS | NA | – | NA | 3.5 mo | female | NS | yes |

Abbreviations: d, day; mo, month; NS, not specified; NA, not applicable; –, negative result; +, positive result
† Study was available as a non-peer-reviewed preprint and had not been published in a refereed journal.
<sup>1</sup> Study presented data from 9 women, but only presented data related to milk produced by 6 women.
<sup>2</sup> Gestational age upon admission.
<sup>3</sup> The infant's breastfeeding status was not specified in the report, but it is presumed that he was at least partially breastfed as the mother was producing milk at 6 mo postpartum.
<sup>4</sup> Study presented data from 3 women but only presented data on the milk produced by 2 women.
<sup>5</sup> Study presented data from 19 women but only presented data on the milk produced by 10 women.
<sup>6</sup> Study presented data from 13 women but only presented data on the milk produced by 3 women.
<sup>7</sup> Days are reported from time of admission, not from time of birth as the other days in this column.

Wang and colleagues (2020) described a healthy, 34-yr-old woman who acquired the infection in week 40 of pregnancy. She gave birth to a male infant via cesarean section. The infant and his mother both tested positive for SARS-CoV-2 using pharyngeal swabs within 36 hr of the delivery. The infant was separated from his mother at delivery and fed infant formula for the duration of the period described in the study. The mother’s milk was collected at 36 hr postpartum; it tested negative for SARS-CoV-2 via RT-PCR. No description of the collection or testing methods was provided. The authors stated they recommended that the mother not breastfeed, but instead pump milk to avoid mastitis.

In another case series from China, Fan and colleagues (2020) described two women who became infected during the third trimester of pregnancy. Patient 1 was 34 yr old and in week 37 of gestation at the time of diagnosis via RT-PCR analysis of a nasopharyngeal swab. She delivered a female infant via cesarean section 6 d after testing positive for SARS-CoV-2 via nasopharyngeal swab. The infant was separated from the mother immediately after delivery, and serial tests of the infant’s nasopharyngeal swabs were negative. A milk sample was collected within 24 hr of delivery and 16 d later; both were negative for SARS-CoV-2. Patient 2 was 29 yr old and in week 36 of gestation at the time of diagnosis via RT-PCR analysis of a nasopharyngeal swab. Her infant was delivered 5 d after she was diagnosed. A single milk sample was collected within 24 hr of delivery; it tested negative for SARS-CoV-2. The authors of this report did not specify how the sample was collected, other than “breastmilk was obtained after the first lactation.”

Chen and colleagues (2020) described milk produced by 6 women infected during pregnancy. The women were 26-34 yr old and between 36 wk 2 d and 39 wk 4 d of gestation at diagnosis. The authors did not provide details on the methods used for milk collection other than “breastmilk samples from patients with COVID-19 pneumonia were collected after their first lactation” and that milk was collected following WHO guidelines, but they did not provide a citation for this collection method. All milk tested negative for the virus, but no information was provided on the methods used for analysis.

In a report by Weiyong Liu and colleagues (2020a) milk produced by two women was tested. One woman was 34 yr old and at 40 wk gestation tested positive for COVID-19 via oropharyngeal swab. Milk was collected and tested at d 1, 2, and 12 postpartum; all samples were negative. Her male infant was delivered via cesarean and tested for SARS-CoV-2 via oropharyngeal swab when he was 1 and 7 days old; both swabs were negative. The other woman was 30 yr old and delivered an infant vaginally after testing positive for SARS-CoV-2. Her infant tested negative at birth using an oropharyngeal swab; milk was collected on d 2 postpartum, it was also negative. No details were provided for methods of collection or analysis.

In a case series by Wei Liu and colleagues (2020b), milk produced by 10 women infected during late pregnancy was tested via RT-PCR for SARS-CoV-2; all samples tested negative. It is noteworthy that this report included data from 19 women, but milk was collected from only 10 of them. The authors did not specify for which of the women milk was collected. None of the 19 infants reported in this study tested positive for SARS-CoV-2 via RT-PCR. The only detail available on collection or testing methods is that RT-PCR was used to test the samples and that “milk was collected after the first lactation.” Despite their results, the authors concluded that delivery should occur in an isolation room, and that infants be separated from infected mothers.

Li and colleagues (2020) described a 30-yr-old woman at 35 wk gestation who was positive for SARS-CoV-2 and who delivered a male infant via emergency cesarean section. The infant was tested immediately upon delivery via oropharyngeal swab, which was negative. After delivery, the infant was kept in isolation away from his mother. Milk was collected immediately after delivery and on d 2 and 3 postpartum; all samples were negative. Again, no information on the collection or testing methods for the milk sample is available in this report.

Dong and colleagues (2020) described a 29-yr-old woman at 34 wk of gestation diagnosed with COVID-19 via nasopharyngeal swab. Nearly a month later, the woman delivered a female infant via cesarean section. The infant was immediately separated from the mother with no contact. The infant consistently tested negative for SARS-CoV-2 via nasopharyngeal swab over the first 12 d of life. However, a blood sample at 2 hr of age was positive for IgG and IgM antibodies to SARS-CoV-2. A milk sample was collected from the mother at d 6 postpartum; it tested negative for SARS-CoV-2 but was not tested for antibodies. No information on the collection or testing methods for the milk sample is included in the report.

In another case study, Yu and colleagues (2020; non-peer-reviewed preprint) described a 32-yr-old woman with a 13-mo-old breastfed male infant. Both the woman and her infant developed symptoms 2 wk after exposure to infected family members and tested positive for COVID-19 2 d after hospital admission. The woman insisted that she remain with her infant during the hospital stay, and the infant continued to breastfeed. Milk samples were collected and analyzed for SARS-CoV-2 on d 1, 8, 15, and 18 after admission, and all tested negative. Milk samples collected on d 8 and d 24 were tested for SARS-CoV-2 specific antibodies. In both samples, the authors identified IgG but not IgM. No details on method of milk collection, SARS-CoV-2 testing, or antibody testing are provided in this report.

Wu and colleagues (2020; non-peer-reviewed preprint) are the only researchers to date who have reported a positive result for SARS-CoV-2 in a human milk sample. All three women (27, 28, and 29 yr old) they studied were infected during the third trimester of pregnancy and delivered infants via cesarean. Milk was collected from each woman on d 1, 6, and 27 postpartum into sterile containers after cleaning the breast with iodine; milk samples were tested via RT-PCR for SARS-CoV-2. Whereas most samples were negative for the virus, the sample collected on d 1 from the 29-yr-old patient was positive. Subsequent testing of milk from this same subject two days later was negative for the virus. No details on the analytical methods used were provided. The infants were also tested for SARS-CoV-2 via both throat and/or anal swabs when they were 1 and/or 3 d old; all were negative.

While the previous reports focused on infected women, there are also three case studies focused on infected infants. In these studies, milk produced by the infants’ mothers was tested for SARS-CoV-2. The youngest of these infants was reported by Cui and colleagues (2020). After being exposed to infected family members, the 55 d old female was admitted to the hospital with symptoms of COVID-19 and diagnosed based on clinical data and exposure history. The infant was “mixed fed.” Her mother’s milk was collected on the first 3 consecutive days of her hospitalization; all were negative for SARS-CoV-2. No information on the collection or testing methods for the milk sample is included in this report. Yuehua and colleagues (2020) reported on a 3-mo-old, breastfed female who was hospitalized and tested via throat swab for SARS-CoV-2; the swab was positive. A single milk sample was collected from the infant’s mother; it tested negative. The authors provided no information on the collection or testing methods for the milk. Importantly, this infant developed symptoms of COVID-19 7 d before her parents became ill. As such, one possibility is that she was infected first and passed the infection to them. Another case report on a mature milk sample comes from Singapore (Kam et al., 2020). This report is particularly interesting as the infant had no symptoms but was hospitalized and tested because his caregivers were all hospitalized with COVID-19 and there was no one to care for him. The infant was 6 mo old and presumably at least partially human milk fed as a sample of milk was successfully collected from his mother. Despite being asymptomatic, a nasopharyngeal swab taken from the infant was positive for SARS-CoV-2. The authors reported that milk produced by the mother on a single day tested negative for the virus but did not specify how many samples were taken. This report provided no data on the methods used for the collection and analysis of these sample(s).

## DISCUSSION

Despite the devastating clinical manifestations of MERS-CoV, SARS-CoV, and SARS-CoV-2, there remains much to be learned about their modes of transmission. Respiratory droplets are a documented source of the virus (World Health Organization, 2020b), but other sources such as breastfeeding and/or human milk may exist. The primary purpose of this review was to examine the evidence (or lack, thereof) for the vertical transmission of SARS-CoV-2 from mother to infant via breastfeeding considering what is known about other human coronaviruses. We also examined the evidence presented in the same reports related to maternal/infant antibody production to the virus.

In total, we identified 13 studies that had tested human milk for human coronaviruses directly. Twelve of these studies were newly published reports on SARS-CoV-2 and human milk, which collectively encompassed 46 milk samples. All but one of these samples tested negative for SARS-CoV-2, and that result was reported in a non-peer-reviewed, on-line preprint. We identified no comparable data for MERS; a single case report for SARS, which yielded a negative result for the presence of the virus but positive results for antibodies specific to SARS-CoV; and no reports of human milk tested for other human coronaviruses. There was one report of antibody tests in milk specific to SARS-CoV-2, which identified IgG but not IgM (Yu et al., 2020). This dearth of high-quality evidence substantially compromises the ability to effectively respond to this pandemic and provide guidance to some of the most vulnerable individuals: pregnant and lactating women and infants.

Limited and weak data suggest MERS may be present in camel milk, but the relevance to SARS-CoV-2 in human milk is unclear. Notably, Reusken and colleagues (2014) reported that milk analyzed in the camelid studies was not collected aseptically; rather, samples were obtained according to local milking customs. As such, it is possible that the presence of MERS-CoV in camel milk could be due to contamination from the milker, the calf, or the environment, rather than milk representing an endogenous source of the virus. This is likely an issue with all the studies on SARS-CoV-2, where only one reported cleaning of the breast prior to sample collection (Wu et al., 2020). However, the limited data available on all three of these viruses (and human coronaviruses, in general) leave many questions unanswered with respect to the role, if any, of human milk in vertical transmission of coronaviruses.

### Box 2.

**KEY POINTS OF ASSAY VALIDATION**

Some factors to consider when validating methods for human milk testing of coronaviruses.

- Method of milk collection: use of manual milk expression vs. electric pump; cleaning procedures of breast and pump; partial vs. full breast expression; foremilk vs. hindmilk.
- Sample handling and storage: container material; temperature; duration of refrigeration/freezing.
- Assay validation: nucleic acid extraction protocols; amplification protocols; reagent selection; proper positive and negative controls; fresh vs. frozen milk.
- Viral quantification and viability: infectious dose; biologically relevant concentrations.

One possible reason that most of the RT-PCR results for the milk samples tested were negative is that the methods used were neither designed nor validated for human milk. Milk is a complex matrix containing substantial fat, DNases (Babina, Kanyshkova, Buneva, & Nevinsky, 2004), RNases (McCormick, Larson, & Rich, 1974; Das, Padhy, Koshy, Sirsat, & Rich, 1976; Ramaswamy, Swamy, & Das, 1993) and other PCR inhibitors (Abu Al-Soud, Jonsson, & Radstrom, 2000; Al-Soud & Radstrom, 2001; Schrader, Schielke, Ellerbroek, & Johne, 2012). Of note is the fact that commonly used silica column-based RNA isolation methods are designed for a limited sample volume, and as such are not suitable for more voluminous liquid samples. Thus, validation of methods using human milk is needed (see **Box 2**). In addition, other than general statements about the timing of collection (e.g., “milk was collected after the first lactation”) and brief descriptions of the RT-PCR assays used for nasal and throat swabs, none of the studies to date has described the methods of collection or how the milk was handled and stored in any detail. In addition, nothing is known about stability of SARS-CoV-2, if present, in human milk and how quickly (or at what temperature) it must be frozen to preserve fidelity. Information on sample collection, handling, and storage is critical to evaluating whether the negative results described in these studies could be due to inadequate methods used.

Another possibility is that there is low abundance of the virus in human milk, and it is often not captured in the limited samples tested so far. For example, in the report on other human coronaviruses by Gagneur and colleagues (2008) 159 maternal-infant dyads were tested (including 161 infants, two sets of twins). In this report, 229E was present in both maternal and infant samples in only 2 dyads. Additionally, in the milk of dromedary camels, MERS-CoV appears to be present at very low abundance (Reusken et al., 2014). This suggests the possibility that a very low viral load in milk might also lead to an inflation of false negatives. Limited evidence from 2 patients suggests that SARS-CoV-2 shedding in respiratory samples peaks at ∼6 d after onset of symptoms (Pan et al., 2020), indicating that timing of sample collection also plays an important role in virus detection.

Only one study has investigated antibodies in milk specific to SARS-CoV-2 (Yu et al. 2020; non-peer-reviewed preprint), and these researchers identified IgG in one milk sample produced by a woman at 13 mo postpartum. While limited to a single study, this finding combined with a large body of literature documenting targeted antibodies in human milk indicate that there may be a protective effect of breastfeeding when the mother is COVID-19 positive. The infant in this study was older than all the other infants described here, was likely not exclusively breastfed (based on reported age), and likely had a more mature immune system than the youngest infants described in other reports. Still, further investigation into this finding is a critical next step in understanding how breastfeeding and/or the infant’s consumption of the complex milieu of human milk impact the infant’s immune response to and clinical manifestations of SARS-CoV-2 infection. This information is an important component in the risk/benefit analysis of developing evidence-based breastfeeding recommendations related to maternal coronavirus infection.

Considering the observations by Yu and colleagues (2020; non-peer-reviewed preprint), other immune protective components of human milk should also be more thoroughly evaluated. While the methods used to test this milk were not fully described, this observation could have impacts on the clinical management of infants born to women diagnosed with COVID-19 during pregnancy and/or lactation. This observation is also supported by the findings of Dong and colleagues (2020) and Zeng and colleagues (2020) who reported that both IgG and IgM antibodies to SARS-CoV-2 were present in the serum of an infant within 2 hr of age, despite multiple negative RT-PCR tests of nasopharyngeal swabs over the first days of life. The presence of circulating antibodies at such an early stage of life could indicate transfer of SARS-CoV-2-specific antibodies from mother to infant during gestation. However, it is noteworthy that IgM antibodies present in the serum of SARS-CoV-2 negative infants are not likely to have originated from the mother during gestation as IgM cannot cross the placental barrier due to size. From the limited data on SARS-CoV, it appears that the presence of antibodies in milk could be influenced by timing of infection, where antibodies to SARS-CoV were detected only in milk produced by a woman who acquired the infection later in pregnancy (Robertson et al., 2004). Together, these observations suggest infant infection may occur *in utero*, but that the virus may simply be absent from the upper respiratory tract immediately after birth and therefore, undetectable on pharyngeal swabs.

Very recent work has demonstrated that, like SARS-CoV and human coronavirus NL63 (Hofmann et al., 2005), angiotensin-converting enzyme 2 (ACE2) is one of the receptors used by SARS-CoV-2 to enter host cells (W. H. Li et al., 2003; Yan et al., 2020). ACE2 is expressed across many body sites and tissue types, including the oral cavity (e.g., tongue and oral mucosa) and in mammary tissue (H. Xu et al., 2020a). If mammary epithelial cells express this receptor, then it follows that viable virus could exist in milk. If it does, then the introduction of virus-containing human milk could represent a mechanism of entry for SARS-CoV-2 and COVID-19 infection for infants.

Another observation worth considering is that, in at least one of the reports (Yuehua et al., 2020), the infant was infected and symptomatic 7 d prior to the infant’s parents. This suggests the possibility that a “reverse” vertical transmission from infant to mother could occur, a phenomenon which has been observed for other pathogens, such as HIV (Belitsky, 1989; Little et al., 2012) and Ebola virus (Sissoko et al., 2016). One possible mechanism for maternal infection in this case is through retrograde flow, where milk and saliva move back into the mammary gland from the infant’s mouth during suckling (Ramsay, Kent, Owens, & Hartmann, 2004). While this mechanism is speculative, it represents a possible route whereby an infant could theoretically transfer a pathogen it has encountered in the environment to the mother. It is also possible that maternal infection could occur through other mechanisms, such as infant respiratory droplets (World Health Organization, 2020) or via fecal matter (Y. Xu et al., 2020b).

### Box 3.

**FUTURE NEEDS**

To understand the role of human milk and SARS-CoV-2 infection, the following points must be rapidly addressed.

- Optimization of human milk collection and storage protocols for SARS-CoV-2 research.
- Validation of assays for identification of SARS-CoV-2 RNA and SARS-CoV-2-specific immune components in human milk.
- Multinational population studies documenting presence or absence of SARS-CoV-2 virus and immune factors (including antibodies) in milk produced by infected women, women with infected infants, and women who have been exposed to SARS-CoV-2; if the virus is identified in milk its viability must be verified.
- Multinational population studies documenting (or not documenting) risk of COVID-19 infections in breastfed vs. non-breastfed infants whose mothers are COVID-19 positive.
- Research delineating implications of skin-to-skin breastfeeding vs. consumption of pumped human milk.

To date, all reports on SARS-CoV-2 and human milk have originated in Asia, specifically China and Singapore. While this limited geography makes sense given the fact that the initial epicenter of this pandemic was in this region, studies from other globally representative populations are needed to make definitive conclusions regarding the possible presence and/or role of SARS-CoV-2 in human milk. Additionally, the importance of a coordinated, international effort by scientists, clinicians, and public health officials to elucidate answers to the many remaining questions related to SARS-CoV-2 and breastfeeding cannot be overemphasized.

## CONCLUSIONS

Human milk is the gold standard for infant feeding. However, confidence as to its safety and best practices around breastfeeding during maternal COVID-19 infection has been compromised by the lack of rigorous evidence as to whether SARS-COV-2 can be vertically transmitted in milk and/or during breastfeeding. As such, there exists an immediate need to rapidly generate rigorous evidence for the role (if any) of human milk and breastfeeding in vertical transmission of COVID-19 from mothers to infants. To accomplish this, validation of analytical methods for the human milk matrix, viability testing, and evaluation of other immune components in milk will all be critical to this effort, especially given the known protective effects of breastfeeding in other infant respiratory infections (**Box 3**; Chantry, Howard, & Auinger, 2006; Duijts, Jaddoe, Hofman, & Moll, 2010). Substantial interdisciplinary research on this topic is required and should be performed rigorously and rapidly to best inform policies regarding early feeding choices and clinical management of breastfeeding mothers infected with SARS-CoV-2 and their infants.

## Data Availability

This manuscript represents a review of existing literature, no data has been generated.

## REFERENCES

Abu Al-Soud, W., Jonsson, L. J., & Radstrom, P. (2000). Identification and characterization of immunoglobulin G in blood as a major inhibitor of diagnostic PCR. Journal of Clinical Microbiology, 38(1), 345–350.

Al-Soud, W. A., & Radstrom, P. (2001). Purification and characterization of PCR-inhibitory components in blood cells. Journal of Clinical Microbiology, 39(2), 485–493. http://doi.org/10.1128/JCM.39.2.485-493.2001

American Academy of Pediatrics. (2020). Initial guidance: management of infants born to mothers with COVID-19. Retrieved April 17, 2020, from https://downloads.aap.org/AAP/PDF/COVID%2019%20Initial%20Newborn%20Guidance.pdf

American Academy of Pediatrics. (2012). Policy statement: breastfeeding and the use of human milk. Pediatrics, 129(3), e827–41. http://doi.org/10.1542/peds.2011-3552

Babina, S. E., Kanyshkova, T. G., Buneva, V. N., & Nevinsky, G. A. (2004). Lactoferrin is the major deoxyribonuclease of human milk. Biochemistry (Moscow), 69(9), 1006–1015. http://doi.org/10.1023/b:biry.0000043543.21217.b3

Belitsky, V. (1989). Children infect mothers in aids outbreak at a soviet hospital. Nature, 337(6207), 493– 493. http://doi.org/10.1038/337493a0

Black, R. F. (1996). Transmission of HIV-1 in the breast-feeding process. Journal of the American Dietetic Association, 96(3), 267–274. http://doi.org/10.1016/S0002-8223(96)00079-X

Boostani, R., Sadeghi, R., Sabouri, A., & Ghabeli-Juibary, A. (2018). Human T-lymphotropic virus type I and breastfeeding; systematic review and meta-analysis of the literature. Iranian Journal of Neurology, 17(4), 174–179.

Centers for Disease Control and Prevention. (2019a). Contraindications to breastfeeding or feeding expressed breast milk to infants. http://doi.org/10.1089/bfm.2016.29002.pjb

Centers for Disease Control and Prevention, Prevention. (2019b). Cytomegalovirus (CMV) and congenital CMV infection. Retrieved April 3, 2020, from https://www.cdc.gov/cmv/clinical/overview.html

Centers for Disease Control and Prevention. (2020a). Coronavirus disease (COVID-19) and breastfeeding. Retrieved April 3, 2020, from https://www.cdc.gov/breastfeeding/breastfeeding-special-circumstances/maternal-or-infant-illnesses/covid-19-and-breastfeeding.html

Centers for Disease Control and Prevention. (2020b). Human immunodeficiency virus (HIV). Retrieved April 3, 2020, from https://www.cdc.gov/breastfeeding/breastfeeding-special-circumstances/maternal-or-infant-illnesses/hiv.html

Chantry, C. J., Howard, C. R., & Auinger, P. (2006). Full breastfeeding duration and associated decrease in respiratory tract infection in US children. Pediatrics, 117(2), 425–432. http://doi.org/10.1542/peds.2004-2283

Chen, H., Guo, J., Wang, C., Luo, F., Yu, X., Zhang, W., … Zhang, Y. (2020). Clinical characteristics and intrauterine vertical transmission potential of COVID-19 infection in nine pregnant women: a retrospective review of medical records. Lancet (London, England), 395(10226), 809–815. http://doi.org/10.1016/S0140-6736(20)30360-3

Cheng, P. K. C., Wong, D. A., Tong, L. K. L., Ip, S.-M., Lo, A. C. T., Lau, C.-S., … Lim, W. (2004). Viral shedding patterns of coronavirus in patients with probable severe acute respiratory syndrome. Lancet (London, England), 363(9422), 1699–1700. http://doi.org/10.1016/S0140-6736(04)16255-7

Conzade, R., Grant, R., Malik, M., Elkholy, A., Elhakim, M., Samhouri, D., … Van Kerkhove, M. (2018). Reported direct and indirect contact with dromedary camels among laboratory-confirmed MERS-CoV cases. Viruses, 10(8), 425–10. http://doi.org/10.3390/v10080425

Coutsoudis, A., Pillay, K., Kuhn, L., Spooner, E., Tsai, W. Y., Coovadia, H. M., South African Vitamin A Study Group. (2001). Method of feeding and transmission of HIV-1 from mothers to children by 15 months of age: prospective cohort study from Durban, South Africa. AIDS (London, England), 15(3), 379–387. http://doi.org/10.1097/00002030-200102160-00011

Cui, Y., Tian, M., Huang, D., Wang, X., Huang, Y., Fan, L., Zha, Y. (2020). A 55-day-old female infant infected with 2019 novel coronavirus disease: presenting with pneumonia, liver injury, and heart damage. The Journal of Infectious Diseases, 376, 584–7. http://doi.org/10.1093/infdis/jiaa113

Das, M. R., Padhy, L. C., Koshy, R., Sirsat, S. M., & Rich, M. A. (1976). Human milk samples from different ethnic-groups contain RNase that inhibits, and plasma-membrane that stimulates, reverse transcription. Nature, 262(5571), 802–805. http://doi.org/10.1038/262802a0

Davis, N. L., Miller, W. C., Hudgens, M. G., Chasela, C. S., Sichali, D., Kayira, D., the Ban study team. (2016). Maternal and breastmilk viral load: impacts of adherence on peripartum HIV infections averted-the breastfeeding, antiretrovirals, and nutrition study. Journal of Acquired Immune Deficiency Syndromes (1999), 73(5), 572–580. http://doi.org/10.1097/QAI.0000000000001145

Dong, L., Tian, J., He, S., Zhu, C., Wang, J., Liu, C., & Yang, J. (2020). Possible vertical transmission of SARS-CoV-2 from an infected mother to her newborn. Jama, 1–3. http://doi.org/10.1001/jama.2020.4621

Duijts, L., Jaddoe, V. W. V., Hofman, A., & Moll, H. A. (2010). Prolonged and exclusive breastfeeding reduces the risk of infectious diseases in infancy. Pediatrics, 126(1), e18–e25. http://doi.org/10.1542/peds.2008-3256

Dunn, D. T., Newell, M. L., Ades, A. E., & Peckham, C. S. (1992). Risk of human immunodeficiency virus type 1 transmission through breastfeeding. The Lancet, 340(8819), 585–588. http://doi.org/10.1016/0140-6736(92)92115-v

Dworsky, M., Yow, M., Stagno, S., Pass, R. F., & Alford, C. (1983). Cytomegalovirus infection of breast milk and transmission in infancy. Pediatrics, 72(3), 295–299.

Fan, C., Lei, D., Fang, C., Li, C., Wang, M., Liu, Y., Wang, S. (2020). Perinatal transmission of COVID-19 associated SARS-CoV-2: should we worry? Clinical Infectious Diseases, ciaa226. http://doi.org/10.1093/cid/ciaa226

Fischer, C., Meylan, P., Graz, M. B., Gudinchet, F., Vaudaux, B., Berger, C., & Roth-Kleiner, M. (2010). Severe postnatally acquired cytomegalovirus infection presenting with colitis, pneumonitis and sepsis-like syndrome in an extremely low birthweight infant. Neonatology, 97(4), 339–345. http://doi.org/10.1159/000260137

Gagneur, A., Dirson, E., Audebert, S., Vallet, S., Legrand-Quillien, M. C., Laurent, Y., Payan, C. (2008). Materno-fetal transmission of human coronaviruses: a prospective pilot study. European Journal of Clinical Microbiology & Infectious Diseases, 27(9), 863–866. http://doi.org/10.1007/s10096-008-0505-7

Hamprecht, K., & Goelz, R. (2017). Postnatal cytomegalovirus infection through human milk in preterm infants: transmission, clinical presentation, and prevention. Clinics in Perinatology, 44(1), 121–130. http://doi.org/10.1016/j.clp.2016.11.012

Hemida, M. G., Elmoslemany, A., Al-Hizab, F., Alnaeem, A., Almathen, F., Faye, B., Peiris, M. (2015). Dromedary camels and the transmission of middle east respiratory syndrome coronavirus (MERS-CoV). Transboundary and Emerging Diseases, 64(2), 344–353. http://doi.org/10.1111/tbed.12401

Hofmann, H., Pyrc, K., van der Hoek, L., Geier, M., Berkhout, B., & Pohlmann, S. (2005). Human coronavirus NL63 employs the severe acute respiratory syndrome coronavirus receptor for cellular entry. Pnas, 102(22), 7988–7993. http://doi.org/10.1073/pnas.0409465102

Iliff, P. J., Piwoz, E. G., Tavengwa, N. V., Zunguza, C. D., Marinda, E. T., Nathoo, K. J., Humphrey, J. H. (2005). Early exclusive breastfeeding reduces the risk of postnatal HIV-1 transmission and increases HIV-free survival. AIDS, 19(7), 699–708. http://doi.org/10.1097/01.aids.0000166093.16446.c9

Jeong, S. Y., Sung, S. I., Sung, J.-H., Ahn, S. Y., Kang, E.-S., Chang, Y. S., Kim, J. (2017). MERS-CoV infection in a pregnant woman in Korea. Journal of Korean Medical Science, 32(10), 1717–1720. http://doi.org/10.3346/jkms.2017.32.10.1717

Jones, C. A. (2001). Maternal transmission of infectious pathogens in breast milk. J. Paediatric Child Health, 37, 576–582. https://doi.org/10.1046/j.1440-1754.2001.00743.x

Kam, K.-Q., Yung, C. F., Cui, L., Lin Tzer Pin, R., Mak, T. M., Maiwald, M., Thoon, K. C. (2020). A well infant with coronavirus disease 2019 (COVID-19) with high viral load. Clinical Infectious Diseases, 361, 1701. http://doi.org/10.1093/cid/ciaa201

Kim, J.-M., Chung, Y.-S., Jo, H. J., Lee, N.-J., Kim, M. S., Woo, S. H., Han, M-G. (2020). Identification of coronavirus isolated from a patient in Korea with COVID-19. Osong Public Health and Research Perspectives, 11(1), 3–7. http://doi.org/10.24171/j.phrp.2020.11.1.02

Lanzieri, T. M., Dollard, S. C., Josephson, C. D., Schmid, D. S., & Bialek, S. R. (2013). Breast milk-acquired cytomegalovirus infection and disease in VLBW and premature infants. Pediatrics, 131(6), e1937–e1945. http://doi.org/10.1542/peds.2013-0076

Lawrence, R. M., & Lawrence, R. A. (2004). Breast milk and infection. Clinics in Perinatology, 31(3), 501–528. http://doi.org/10.1016/j.clp.2004.03.019

Li, W. H., Moore, M. J., Vasilieva, N., Sui, J. H., Wong, S. K., Berne, M. A., Farzan, M. (2003). Angiotensin-converting enzyme 2 is a functional receptor for the SARS coronavirus. Nature, 426(6965), 450–454. http://doi.org/10.1038/nature02145

Li, Y., Zhao, R., Zheng, S., Chen, X., Wang, J., Sheng, X., Sheng, J. (2020). Lack of vertical transmission of severe acute respiratory syndrome coronavirus 2, China. Emerging Infectious Diseases, 26(6), 727. http://doi.org/10.3201/eid2606.200287

Little, K. M., Kilmarx, P. H., Taylor, A. W., Rose, C. E., Rivadeneira, E. D., & Nesheim, S. R. (2012). A review of evidence for transmission of HIV from children to breastfeeding women and implications for prevention. The Pediatric Infectious Disease Journal, 31(9), 938–942. http://doi.org/10.1097/INF.0b013e318261130f

Liu, Weiyong., Wang, Q., Zhang, Q., Chen, L., Chen, J., & Zhang, B. (2020a). Coronavirus disease 2019 (COVID-19) during pregnancy: A case series. Preprints, 2020020373.

Liu, Wei, Wang, J., Li, W., Zhou, Z., Liu, S., & Rong, Z. (2020b). Clinical characteristics of 19 neonates born to mothers with COVID-19. Frontiers in Medicine, doi: 10.1007/s11684-020-0772-y.

Mahase, E. (2020). Coronavirus covid-19 has killed more people than SARS and MERS combined, despite lower case fatality rate. BMJ, 368, m641. http://doi.org/10.1136/bmj.m641

McCormick, J. J., Larson, L. J., & Rich, M. A. (1974). Rnase inhibition of reverse-transcriptase activity in human milk. Nature, 251(5477), 737–740. http://doi.org/10.1038/251737a0

Memish, Z. A., Cotten, M., Meyer, B., Watson, S. J., Alsahafi, A. J., Rabeeah Al, A. A., Drosten, C. (2014). Human infection with MERS coronavirus after exposure to infected camels, Saudi Arabia, 2013. Emerging Infectious Diseases, 20(6), 1012–1015. http://doi.org/10.3201/eid2006.140402

Minamishima, I., Ueda, K., Minematsu, T., Minamishima, Y., Umemoto, M., Take, H., & Kuraya, K. (1994). Role of breast-milk in acquisition of cytomegalovirus-infection. Microbiology and Immunology, 38(7), 549–552. http://doi.org/10.1111/j.1348-0421.1994.tb01821.x

Moriuchi, H., Masuzaki, H., Doi, H., & Katamine, S. (2013). Mother-to-child transmission of human T-cell lymphotropic virus type 1. The Pediatric Infectious Disease Journal, 32(2), 175–177. http://doi.org/10.1097/INF.0b013e31827efc39

Nduati, R., Richardson, B. A., John, G., Mbori-Ngacha, D., Mwatha, A., Ndinya-Achola, J., Kreiss, J. (2001). Effect of breastfeeding on mortality among HIV-1 infected women: a randomized trial. The Lancet, 357(9269), 1651–1655. http://doi.org/10.1016/S0140-6736(00)04820-0

Pan, Y., Zhang, D., Yang, P., Poon. LLM., Wang, Q. (2020). Viral load of SARS-CoV-2 in clinical samples. Lancet Infect Dis, 20(4): 411–412. http://doi.org/10.1016/S1473-3099(20)30113-4.

Ramaswamy, H., Swamy, C., & Das, M. R. (1993). Purification and characterization of a high-molecular-weight ribonuclease from human-milk. Journal of Biological Chemistry, 268(6), 4181–4187.

Ramsay, D. T., Kent, J. C., Owens, R. A., & Hartmann, P. E. (2004). Ultrasound imaging of milk ejection in the breast of lactating women. Pediatrics, 113(2), 361–367. http://doi.org/10.1542/peds.113.2.361

Reusken, C. B., Farag, E. A., Jonges, M., Godeke, G. J., El-Sayed, A. M., Pas, S. D., Koopmans, M. P. (2014). Middle East respiratory syndrome coronavirus (MERS-CoV) RNA and neutralizing antibodies in milk collected according to local customs from dromedary camels, Qatar, April 2014. Euro Surveillance: European Communicable Disease Bulletin, 19(23), 1–9. http://doi.org/10.2807/1560-7917.ES2014.19.23.20829

Robertson, C. A., Lowther, S. A., Birch, T., Tan, C., Sorhage, F., Stockman, L., Bresnitz, E. (2004). SARS and pregnancy: A case report. Emerging Infectious Diseases, 10(2), 345–348. http://doi.org/10.3201/eid1002.030736

Rosadas, C., & Taylor, G. P. (2019). Mother-to-child HTLV-1 transmission: unmet research needs. Frontiers in Microbiology, 10, 999. http://doi.org/10.3389/fmicb.2019.00999

Schrader, C., Schielke, A., Ellerbroek, L., & Johne, R. (2012). PCR inhibitors occurrence, properties and removal. Journal of Applied Microbiology, 113(5), 1014–1026. http://doi.org/10.1111/j.1365-2672.2012.05384.x

Schwartz, D. A., & Graham, A. L. (2020). Potential maternal and infant outcomes from coronavirus 2019-nCoV (SARS-CoV-2) infecting pregnant women: lessons from SARS, MERS, and other human coronavirus infections. Viruses, 12(2), 194–16. http://doi.org/10.3390/v12020194

Semba, R. D., Kumwenda, N., Hoover, D. R., Taha, T. E., Quinn, T. C., Mtimavalye, L., Chiphangwi, J. D. (1999). Human immunodeficiency virus load in breast milk, mastitis, and mother-to-child transmission of human immunodeficiency virus type 1. The Journal of Infectious Diseases, 180(1), 93–98. http://doi.org/10.1086/314854

Sissoko, D., Keïta, M., Diallo, B., Aliabadi, N., Fitter, D. L., Dahl, B. A., Duraffour, S. (2016). Ebola virus persistence in breast milk after no reported illness: a likely source of virus transmission from mother to child. Clinical Infectious Diseases, 388, ciw793–4. http://doi.org/10.1093/cid/ciw793

Stagno, S., & Cloud, G. A. (1994). Working parents the impact of day-care and breast-feeding on cytomegalovirus infections in offspring. PNAS, 91(7), 2384–2389. http://doi.org/10.1073/pnas.91.7.2384

Stockman, L. J., Lowther, S. A., Coy, K., Saw, J., & Parashar, U. D. (2004). SARS during pregnancy, United States. Emerging Infectious Diseases, 10(9), 1689–1690. http://doi.org/10.3201/eid1009.040244

Tyrrell, D., & Bynoe, M. L. (1965). Cultivation of a novel type of common-cold virus in organ cultures. BMJ, 1(5448), 1467–1470. http://doi.org/10.1136/bmj.1.5448.1467

United Nations Children’s Fund (UNICEF). (2020). Coronavirus disease (COVID-19): What parents should know. Retrieved April 14, 2020, from https://www.unicef.org/stories/novel-coronavirus-outbreak-what-parents-should-know

van Doremalen, N., Bushmaker, T., Karesh, W. B., & Munster, V. J. (2014). Stability of middle east respiratory syndrome coronavirus in milk. Emerging Infectious Diseases, 20(7), 1263–1264. http://doi.org/10.3201/eid2007.140500

Wang, S., Guo, L., Chen, L., Liu, W., Cao, Y., Zhang, J., & Feng, L. (2020). A case report of neonatal COVID-19 infection in China. Clinical Infectious Diseases, 348, 1953. http://doi.org/10.1093/cid/ciaa225

Willumsen, J. F., Filteau, S. M., Coutsoudis, A., Newell, M.-L., Rollins, N. C., Coovadia, H. M., & Tomkins, A. M. (2003). Breastmilk RNA viral load in HIV-infected South African women: effects of subclinical mastitis and infant feeding. AIDS (London, England), 17(3), 407–414. http://doi.org/10.1097/00002030-200302140-00015

World Health Organization. (2020a). Clinical management of severe acute respiratory infection (SARI) when COVID-19 disease is suspected: interim guidance, Retrieved 13 April 2020, from https://www.who.int/publications-detail/clinical-management-of-severe-acute-respiratory-infection-when-novel-coronavirus-(ncov)-infection-is-suspected

World Health Organization. (2020b). Modes of transmission of virus causing COVID-19: implications for IPC precaution recommendations. Retrieved April 3, 2020, from https://www.who.int/news-room/commentaries/detail/modes-of-transmission-of-virus-causing-covid-19-implications-for-ipc-precaution-recommendations

World Health Organization. (2009). Acceptable medical reasons for use of breast-milk substitutes. Retrieved 15 April 2020, from https://www.who.int/nutrition/publications/infantfeeding/WHO_NMH_NHD_09.01/en/

World Health Organization. (2019). WHO MERS-CoV global summary and assessment of risk. Retrieved 15 April 2020, from https://www.who.int/publications-detail/who-mers-cov-global-summary-and-assessment-of-risk

World Health Organization. (2016). Guideline: Updates on HIV and infant feeding. The duration of breastfeeding, and support from health services to improve feeding practices among mothers living with HIV. Geneva: World Health Organization.

Wu, Y., Liu, C., Dong, L., Zhang, C., Chen, Y., Liu, J., Huang, H-F. (2020). Viral shedding of COVID-19 in pregnant women. SSRN, preprint. http://dx.doi.org/10.2139/ssrn.3562059

Xu, H., Zhong, L., Deng, J., Peng, J., Dan, H., Zeng, X., Chen, Q. (2020a). High expression of ACE2 receptor of 2019-nCoV on the epithelial cells of oral mucosa. International Journal of Oral Science, 12(8). http://doi.org/10.1038/s41368-020-0074-x

Xu, Y., Li, X., Zhu, B., Liang, H., Fang, C., Gong, Y., Gong, S. (2020b). Characteristics of pediatric SARS-CoV-2 infection and potential evidence for persistent fecal viral shedding. Nature Medicine, 395, 1– 4. http://doi.org/10.1038/s41591-020-0817-4

Yan, R., Zhang, Y., Li, Y., Xia, L., Guo, Y., & Zhou, Q. (2020). Structural basis for the recognition of SARS-CoV-2 by full-length human ACE2. Science, 367(6485), 1444–1448. http://doi.org/10.1126/science.abb2762

Yu, Y., Xu, J., Li, Y., Hu, Y., Li, B. (2020). Breast milk-fed infant of COVID-19 pneumonia mother: a case report. Research Square, preprint. http://doi.org/10.21203/rs.3.rs-20792/v1

Yuehua, Z., Daojiong, L., Meifang, X., Jiachong, W., Yong, W., Zhixian, L., Wei, X. (2020). A case of three-month-old infant with new coronavirus infection. Chinese Journal of Pediatrics, 58(3), 182–184.

Ziegler, J. B., Johnson, R. O., Cooper, D. A., & Gold, J. (1985). Postnatal transmission of aids-associated retrovirus from mother to infant. The Lancet, 1(8434), 896–898. http://doi.org/10.1016/s0140-6736(85)91673-3

Zeng, H., Xu, C., Fan, J., Tang, Y., Deng, Q., Zhang, W., Long, X. (2020). Antibodies in infants born to mothers with COVID-19 pneumonia. JAMA. Published online March 26, 2020. http://doi.org/10.1001/jama.2020.4861

